# Delirium and High Sedation Levels are Common in Critically Ill Patients with COVID-19 and Associated with Poor Long-Term Outcomes

**DOI:** 10.1101/2024.12.17.24319167

**Authors:** Abigail A. Bucklin, Wolfgang Ganglberger, Ryan A. Tesh, Syed A. Quadri, Muhammad Abubakar Ayub, Susan S. Maher, Miguel Patino Montoya, Preeti Malik, Haitham S. Alabsi, Jonathan Rosand, Eyal Y. Kimchi, Oluwaseun Akeju, Shibani S. Mukerji, Jeanine Wiener-Kronish, M. Brandon Westover

**Author notes:** Corresponding Author: M. Brandon Westover, MD, PhD, Department of Neurology, Massachusetts General Hospital, 55 Fruit Street, Boston, MA 02114. Co-first authors (Abigail A. Bucklin, Wolfgang Ganglberger, Ryan A. Tesh). Co-senior authors (Jeanine Wiener-Kronish, M. Brandon Westover).

## Abstract

**Background:** We investigated delirium prevalence and potential effects of long-term sedation in critically ill COVID-19 patients; to identify opportunities for improving sedation practices and delirium prevention.

**Methods:** This prospective, single-center, observational cohort study was conducted from April-June 2020. Adult COVID-19 patients were eligible if admitted to an ICU with mechanical ventilation/intravenous sedation; or a general care unit with brain monitoring due to altered mental status. Patients were evaluated daily until discharge using the Richmond Agitation-Sedation Scale, Confusion Assessment Method for the ICU, and CAM-Severity. Cumulative doses of sedation and paralytic medications were recorded. At three months post-enrollment, cognition, mood, and quality of life were measured by the Telephone Interview for Cognitive Status (TICS), Center for Epidemiologic Studies Depression Scale 10-item (CES-10), and EuroQol 5-Dimension-3 Level (EQ-5D-3L), respectively.

**Results:** 67 patients were enrolled, with a mean (SD) age of 59 (12) years, 30 (45%) Hispanic, 43 (64%) developing acute respiratory distress syndrome, 55 (82%) mechanically ventilated (mean duration of 22.9 days), and 5 comatose for the entire study. Of the 62 patients assessed for delirium, 61 (98%) had delirium at least once, with a mean (SD) of 12.7 (13.0) days. >90% of patients received opioids, benzodiazepines, or propofol at least once; median (IQR) total dose of 37.4 (78.9) mg (fentanyl equivalents), 52.5 (813.3) mg (midazolam equivalents), and 46 (53) g (propofol), respectively. At follow-up, 40 (60%) patients were reached, while 16 (24%) were deceased/comfort measures. Patients showed reductions in cognition, mood, and quality of life with median (IQR) scores for TICS (0-41): 30 (26-33); CES-D-10 (0-30): 6 (4-12); EQ-5D-3L (1-3): 2 (mobility, self-care, usual activities, pain/discomfort).

**Conclusion:** Critically and acutely ill patients with COVID-19 early in the pandemic experienced a high rate of delirium and sedation. Large doses of sedatives may contribute to greater delirium burden during hospitalization, and lead to poor clinical outcomes.

## Introduction

Severe acute respiratory syndrome coronavirus 2 (SARS-CoV-2), commonly known as COVID-19, was first reported in December 2019 in Wuhan, China. Just over a year later, there were more than 1 billion confirmed cases and 2 million confirmed deaths globally ^1^. At the start of our study, nearly a quarter of reported cases were in the United States, where over 60,000 people had died from COVID-19 ^2^. As hospitalizations for severe COVID-19 surged, it became evident that patients in intensive care units (ICUs) with COVID-19 faced a particularly high risk of developing delirium. One study found that 69% of ICU patients with COVID-19 experienced agitation, and 65% developed delirium ^3^.

Delirium is an acute neurologic syndrome characterized by confusion, inattention, and a fluctuating course, often accompanied by agitation, lethargy, or both ^4^. Among hospitalized patients, its incidence ranges from 20% to 80% ^5-7^, with critically ill, mechanically ventilated patients at the highest risk, exceeding 90% in some reports ^6,8^. Delirium has led to poorer critical care outcomes, such as prolonged ventilation, increased mortality, and other ICU-related complications. For elderly patients with COVID-19, delirium was associated with a fourfold increase in in-hospital mortality compared to those without delirium ^9^. Additionally, delirium during hospitalization has been linked to long-term cognitive impairments after discharge ^10^.

While prior research highlighted high rates of delirium early in the COVID-19 pandemic ^3,11,12^, the quantities of sedation and narcotics administered and their potential contribution to delirium have not been thoroughly examined. The goal of this study was to use once-daily delirium assessments together with medication records to investigate the potential cognitive effects of long-term sedation in patients with COVID-19. This study documents some of the largest quantities of sedation and narcotics administered to ICU patients during the pandemic. Our findings suggest that sedation and narcotics administration practices during the early stages of the pandemic were problematic, highlighting critical opportunities for improvement.

## Methods

### Study design, setting, and participants

We conducted a single-center, prospectively designed, observational cohort study of adults admitted to Massachusetts General Hospital (MGH) with COVID-19 between April 23 and June 11, 2020. Eligible participants were adults aged 18 and older with laboratory-confirmed COVID-19 who met one of the following criteria: 1) admission to an ICU with either mechanical ventilation or intravenous sedative administration, or 2) admission to a general care unit with brain monitoring via EEG due to concerns about altered mental status. The study design is compatible with STROBE guidelines.

### Standard Protocol Approvals, Registrations, and Patient Consents

This study was conducted as a Clinical Quality Improvement Initiative at MGH to investigate sedation practices in critically ill patients with COVID-19 and the potential impact that sedatives have on delirium. According to the Partners Human Research Committee, it qualified for an exemption from formal oversight by the Mass General Brigham Institutional Review Board and met the criteria for a waiver of informed consent. The study was performed in compliance with the ethical principles outlined in the 1964 Declaration of Helsinki, its subsequent amendments, and comparable ethical standards.

### Clinical Assessments

Patients were assessed once daily at the bedside by study staff from enrollment until hospital discharge. Evaluations included the Richmond Agitation-Sedation Scale (RASS) to measure arousal levels, the Confusion Assessment Method for the ICU (CAM-ICU) to determine the presence of delirium, and the CAM-Severity (CAM-S, Long Form: 0–19) to quantify the severity of delirium symptoms ^13-15^. For non-English speaking, evaluations were performed in the presence of an interpreter. For comatose patients (RASS -4 or -5), CAM-ICU and CAM-S assessments were not conducted. Additionally, medical records were reviewed to collect RASS scores routinely documented by nursing staff from admission onward.

For descriptive purposes, ‘Days delirium’ refers to days with CAM-ICU-defined delirium, based on the presence of two major criteria (acute onset/fluctuating course and inattention) and at least one minor criterion (altered level of consciousness or disorganized thinking). ‘Days delirium or coma’ includes days with CAM-ICU-defined delirium or coma (RASS -4 or -5). ‘Sum CAM-S’ represents the total of all CAM-S scores recorded for each patient. Delirium was further classified into subtypes: hyperactive or hypoactive. Hyperactive delirium was defined by either a RASS >0 or the presence of psychomotor agitation on the CAM-S, while hypoactive delirium was defined by either a RASS <0 or the presence of psychomotor retardation on the CAM-S.

At three months post-enrollment, patients were contacted via telephone to assess for sequelae of critical illness and to identify needs for follow-up medical care. A standardized questionnaire was administered, including: 1) Telephone Interview for Cognitive Status (TICS, 0-41) ^16^ where lower scores represent greater impairments; 2) Center for Epidemiologic Studies Depression Scale 10-item (CES-D-10, 0-30) ^17^ with higher scores representing a more depressed mood; and 3) EuroQol 5-Dimension-3 Level (EQ-5D-3L) ^18^ to assess mobility, self-care, usual activities, pain/discomfort, and anxiety/depression where labels (1-3) describe the different severity levels of a dimension (ranging from no problem to extreme problems). Although the dimensions have no arithmetic properties, we derive a summed total score for descriptive purposes. Patients were offered resources, including clinic visits, to help cope with any problems identified during the interview. Baseline functional status was retrospectively determined by reviewing medical records, using the Lawton Instrumental Activities of Daily Living (IADL, 0-8) scale, where lower scores indicate lower function/dependence) ^19^. This assessment incorporated physician documentation and physical/occupational therapy evaluations when available.

### Medications

The total amount of sedation and paralytic medications administered to patients during hospitalization was collected. Cumulative doses of opioids and benzodiazepines were converted into fentanyl and midazolam equivalents, respectively. Details of the conversion are provided in the supplemental material.

### Exposure Strata

Patients were divided into two groups based on the median number of ‘Days with delirium or coma’: 1) ‘low delirium’ group, defined as ‘Days with delirium or coma’ ≤25; 2) ‘high delirium’ group, defined as ‘Days with delirium or coma’ >25. Delirium prevalence statistics and follow-up scores were computed separately for each group. Statistical significance was assessed using a t-test or a Mann-Whitney U test, with the significance level at p <0.05. Additional groupings were created based on different exposure strata; details on these groupings can be found in the supplemental material.

### Linear Regression

Stepwise multivariable regression was conducted using forward selection with the partial F-test ^20,21^. Follow-up outcomes were treated as dependent variables, while delirium prevalence (the primary exposure of interest) and covariates were treated as independent variables. The regression analysis was performed twice: once with the exposure variable forced to be included first and a second time without this constraint. Further details on the modeling process are provided in the supplemental material.

### Data availability

The data supporting the findings reported in this article, including text, tables, and figures, are available from the corresponding author upon reasonable request.

## Results

### Dataset characteristics

A total of 67 patients were enrolled between April and June 2020. Of these, 55 patients were admitted to one of two COVID ICUs with either mechanical ventilation or intravenous sedative administration, and 12 were admitted to a general care unit with brain monitoring via EEG due to concerns about altered mental status. **Figure S1** shows a flow diagram of subject enrollment. The mean (SD) age was 59 (12) years, and 43 (64%) patients had hypoxemic respiratory failure/ARDS due to COVID-19. Additional demographic data are presented in **Table 1**.

**Table 1:** Demographics.

| **Characteristic** | **Total (n = 67)** |
| --- | --- |
| **Age, years: Mean (SD)** | 59 (12%) |
| **Sex** |  |
| Female: n (%) | 27 (40%) |
| Male: n (%) | 40 (60%) |
| **Race** |  |
| Asian: n (%) | 2 (3%) |
| Black: n (%) | 8 (12%) |
| Native American or other Pacific Islander: n (%) | 0 (0%) |
| White: n (%) | 27 (40%) |
| Other or Unknown: n (%) | 30 (45%) |
| **Ethnicity<sup>b</sup>** |  |
| Hispanic: n (%) | 30 (45%) |
| Non-Hispanic: n (%) | 26 (39%) |
| Unknown: n (%) | 11 (16%) |
| **Primary language** |  |
| English: n (%) | 26 (39%) |
| Spanish: n (%) | 34 (51%) |
| Other or Unknown: n (%) | 7 (10%) |
| **BMI (kg/m<sup>2</sup>): Mean (SD)** | 30.7 (6.8) |
| **Charlson Comorbidity Index (CCI)<sup>46</sup>** |  |
| 0 | 28 (42%) |
| 1-2 | 28 (42%) |
| 3-4 | 8 (12%) |
| ≥ 5 | 3 (4%) |
| **Primary diagnosis** |  |
| COVID-19: n (%) | 12 (18%) |
| Hypoxemic Respiratory Failure/ARDS due to COVID-19: n (%) | 43 (64%) |
| Hypercarbic Respiratory Failure/ARDS due to COVID-19: n (%) | 3 (5%) |
| Altered Mental Status: n (%) | 3 (5%) |
| Other: n (%) | 6 (9%) |
| **Disposition/discharge level of support** |  |
| Home, self-care | 3 (4%) |
| Home, with services | 7 (10%) |
| Subacute Rehab (SAR)/Skilled Nursing Facility (SNF) | 3 (4%) |
| Acute Inpatient Rehabilitation Facility (IRF) | 16 (24%) |
| Long-Term Care (LTC) Facility or Nursing Home | 22 (33%) |
| Hospice Service | 2 (3%) |
| Deceased | 14 (21%) |

Swimmer plots show the status of patients (delirium, coma, or neither) during their hospitalization (**Figure 1**). 5 (7%) patients were comatose for the entire study. Of the 62 patients assessed for delirium, 61 (98%) experienced delirium at least once, with a mean (SD) of 12.7 (13.0) days. The mean (SD) CAM-S score was 8.2 (2.7), and all patients had at least one day with a CAM-S score ≥7 (see **Figure S2** for CAM-S swimmer plots). 40 (65%) patients experienced at least one episode of hyperactive delirium, with a median (IQR) of 2 (4) days. 57 (92%) patients experienced at least one episode of hypoactive delirium, with a median (IQR) of 4 (5) days. 30 (44%) patients were comatose at least once during the study; and 63 (94%) were comatose at least once during hospitalization, when considering RASS scores routinely documented by nursing staff from admission onward (see **Figure S3** for RASS swimmer plots).

**Figure 1:**
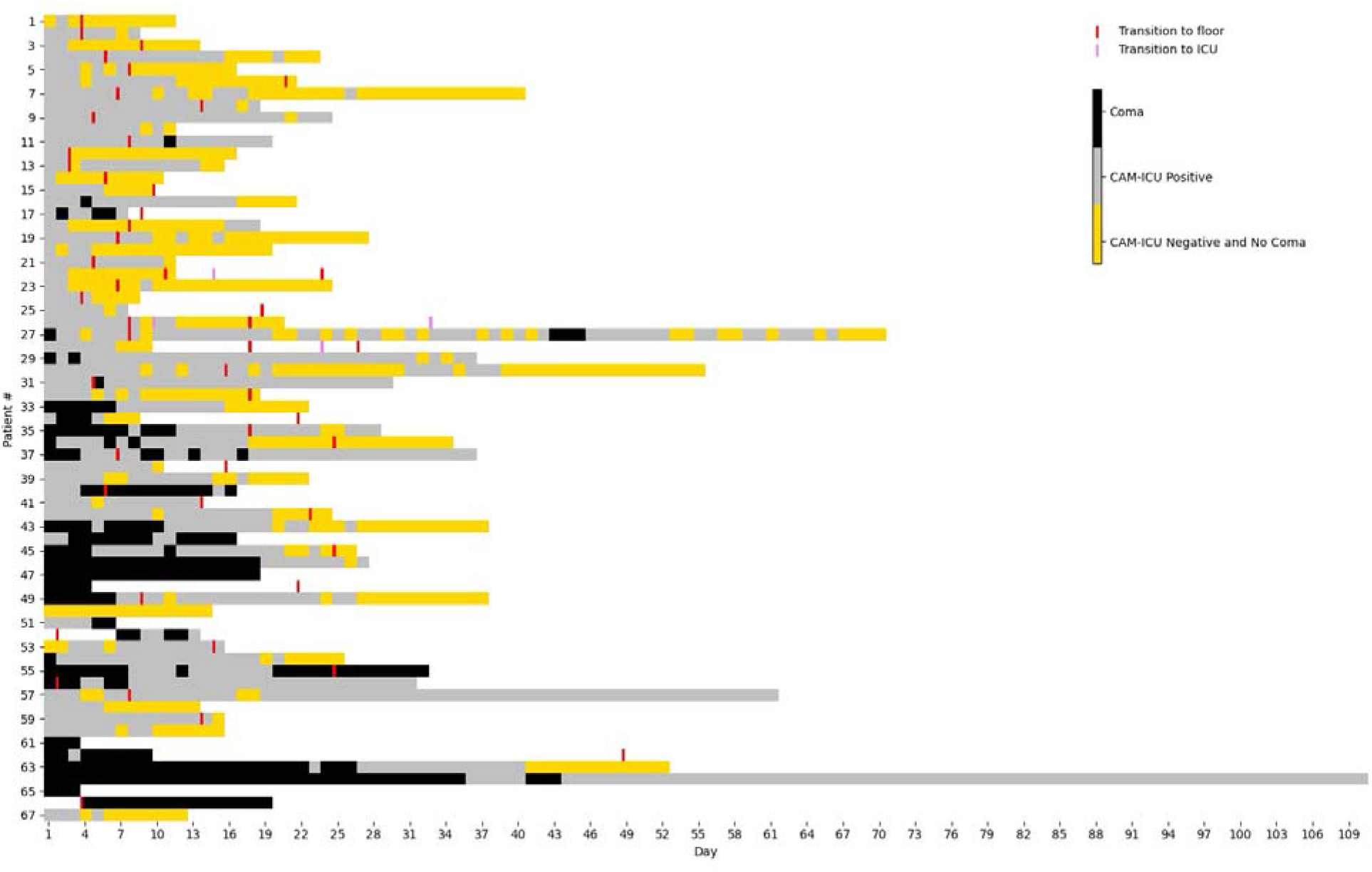
Swimmer plots indicating patients who had delirium (CAM-ICU positive), coma (RASS -4 or - 5), or did not have delirium or coma. Each bin represents one day. Transitions from the ICU to the floor, and from the floor back to the ICU are also shown.

### Follow-up outcomes

Of the 67 patients enrolled, 40 (60%) completed some or all of the 3-month follow-up questionnaire, 11 (16%) could not be reached, and 16 (24%) were deceased or on comfort measures only (CMO). 37 patients completed the TICS [0-41], with a median (IQR) total score of 30 (26-33), or ambiguous for cognitive impairment. Among the 36 patients who completed the CES-D-10 [0-30], median (IQR) scores were 6 (4-12) or a slightly depressed mood, and 12 (33%) patients were considered depressed (with score of 10 or greater). Among the 39 patients who completed the EQ-5D-3L, median (IQR) scores for the five dimensions were: 2 (1-2) or some problems with mobility, self-care, and usual activities; 2 (1-3) or some problems with pain/discomfort, and 1 (1-2) or no problem with anxiety/depression. For descriptive purposes, the median (IQR) for the summed EQ-5D-3L total score was 9 (6-11), or some problems with quality of life. Patients also reported additional post-discharge issues not specifically asked about, including foot pain (5/40; 12.5%), hip or back pain (4/40; 10%), nerve pain (4/40; 10%), chronic pain (4/40; 10%), shoulder/arm pain (3/40; 7.5%), numbness in the extremities (hands/feet) (3/40; 7.5%), weakness (3/40; 7.5%), hair loss (3/40; 7.5%), and throat pain (1/40; 2.5%). In contrast, all but three patients had been independent in daily activities before hospitalization. **Figure 2** provides violin and boxplots summarizing patient scores across the study instruments.

**Figure 2:**
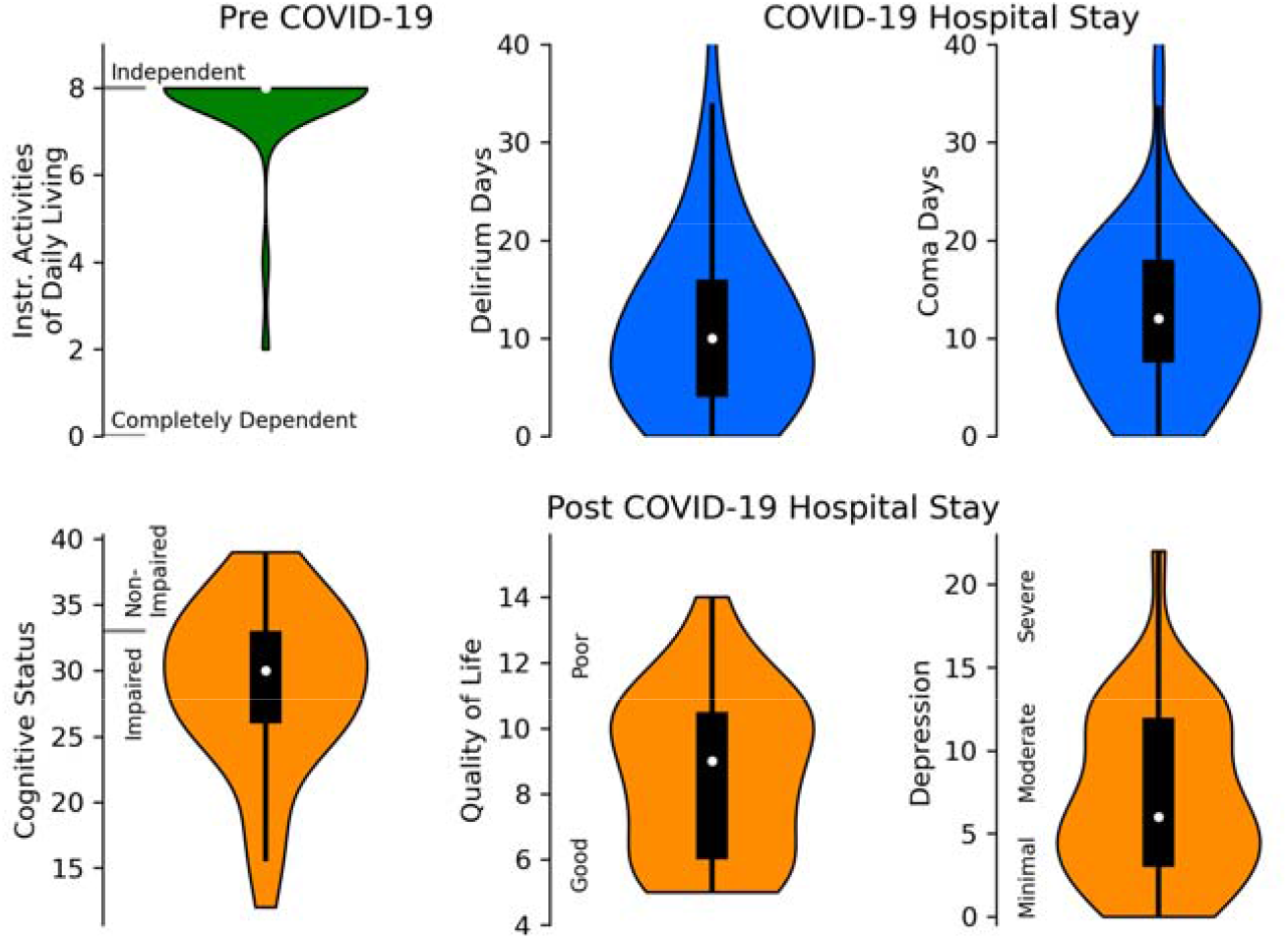
Violin and boxplots showing patient scores across the various instruments used in the study. The white circle in the plots indicates the median score for each metric. Instrumental Activities of Daily Living (IADL, 0-8) (top left) represents patients’ baseline functional status before COVID-19 infection, where lower scores indicate lower function/dependence. Delirium days (top middle) and coma days (top right) reflect patients’ status during hospitalization for COVID-19, as assessed using the Confusion Assessment Method for the ICU (CAM-ICU) and the Richmond Agitation Sedation Scale (RASS), respectively. At the 3-month follow-up, cognition (bottom left) was assessed using the Telephon Interview for Cognitive Status (TICS, 0-41), where suggested qualitative interpretive ranges include a total score of 33-41 (non-impaired), 26-32 (ambiguous), 21-25 (mildly impaired), ≤20 (moderately to severely impaired). Quality of life (bottom middle) was measured with the EuroQol 5-Dimension-3 Level (EQ-5D-3L), which evaluates mobility, self-care, usual activities, pain/discomfort, and anxiety/depression. For descriptive purposes, a higher summed total score indicates more issues in these domains. Depression (bottom right) was measured using the Center for Epidemiologic Studies Depression Scale 10-item (CES-D-10, 0-30), where higher scores represent a more depressed mood (a score of 10 or greater is considered depressed).

### Medications

Patients received various medications during hospitalization, including opioids (64; 96%), benzodiazepines (60; 90%), antipsychotics (60; 90%), propofol (63; 94%), dexmedetomidine (55; 82%), ketamine (31; 46%), isoflurane (11; 16%), and paralytics (60; 90%). Cumulative doses, given as median (IQR), were opioids [37.4 (78.9) mg (fentanyl equivalents)], benzodiazepines [52.5 (813.3) mg (midazolam equivalents) or 26.25 (406.65) mg (lorazepam equivalents)], antipsychotics [245 (722) g], propofol [46 (53) g], dexmedetomidine [6.3 (12.1) mg], ketamine [0.0 (12,291.8) mg]. The mean (SD) duration of isoflurane and paralytic use was 0.4 (1.1) and 6.8 (7.2) days, respectively.

### Exposure Strata

Results of the t-test between the ‘low delirium’ and ‘high delirium’ groups are shown in **Table 2**. Compared to the ‘low delirium’ group, the ‘high delirium’ group exhibited the following significant differences: 1) Longer ICU length of stay: higher mean (SD) of 28.0 (14.6) days vs. 16.8 (8.2) days (p=0.0004); 2) Longer hospital length of stay: higher mean (SD) of 49.8 (21.6) days vs. 26.1 (10.5) days (p<0.0001); 3) Lower TICS scores at 3-months: lower mean (SD) scores of 26.3 (6.6) vs. 31.0 (5.0) (p=0.018); 4) Lower EQ-5D-3L scores at 3-months: lower mean (SD) scores of 9.8 (2.3) vs. 7.7 (2.2) (p=0.0062). The ‘high delirium’ group also received significantly higher amounts of medications, including opioids: median (IQR) of 69.4 (82.0) mg vs. 32.7 (53.6) mg (p=0.0296); benzodiazepines: 136.3 (1517.1) mg vs. 8.0 (265.3) mg (p=0.0072); antipsychotics: 410 (879) g vs. 130 (270) g (p=0.0025); propofol: 54 (39) g vs. 36 (58) g (p=0.0287); ketamine: 1971.6 (18993.5) mg vs. 0.0 (6138.5) mg (p=0.0323). Additionally, the ‘high delirium’ group had a longer median (IQR) duration of paralytic use: 9.5 (9.0) days vs. 4.1 (3.0) days (p=0.0026). For detailed results across different exposure strata, refer to **Tables S1-S3**.

**Table 2:** Exposure Strata.

| | All Patients | Days Delirium or<br>Coma $\leq 25$ | Days Delirium or Coma<br>> 25 | t-statistic (p-value),<br>U-statistic for<br>medications |
| --- | --- | --- | --- | --- |
| **<u>Hospitalization Data</u>** |  |  |  |  |
| N Patients | 62 | 31 | 31 |  |
| Sex |  |  |  |  |
| Male <sup>a</sup> | 40 (60) | 15.5 (50) | 22 (70) | -1.8 (0.0719) |
| Female <sup>a</sup> | 27 (40) | 15.5 (50) | 9 (30) |  |
| Age (years) <sup>b</sup> | 59 (13) | 57 (14) | 60 (12) | -0.8 (0.42) |
| BMI (kg/m<sup>2</sup>) <sup>b</sup> | 30.7 (7.0) | 31.8 (6.5) | 29.5 (7.4) | 1.3 (0.197) |
| ICU Length of Stay (days) <sup>b</sup> | 22.4 (13.0) | 16.8 (8.2) | **28.0 (14.6) | -3.7 (0.0004) |
| Hospital Length of Stay (days) <sup>b</sup> | 38.0 (20.6) | 26.1 (10.5) | **49.8 (21.6) | -5.5 (0.0) |
| Days Intubated <sup>b</sup> | 18.9 (11.4) | 15.6 (6.6) | *23.1 (14.5) | -2.4 (0.0199) |
| Days Intubated or Tracheostomy <sup>b</sup> | 22.9 (13.5) | 17.0 (8.7) | **30.0 (14.9) | -3.8 (0.0004) |
| CCI Score <sup>b</sup> | 1.2 (1.5) | 1.5 (1.5) | 1.0 (1.6) | 1.3 (0.1896) |
| Mortality <sup>a</sup> | 10 (16) | 4 (13) | 6 (19) | -0.7 (0.50) |
| Sequential Organ Failure Assessment<br>(SOFA) score <sup>47 b</sup> | 5.1 (4.0) | 4.9 (4.0) | 5.3 (4.1) | -0.4 (0.6846) |
| Patients with CAM-ICU $\geq 3$ ever<sup>a</sup> | 61 (98) | 30 (97) | 31 (100) | |
| CAM-ICU Score <sup>b</sup> | 2.7 (1.0) | 2.3 (1.1) | 3.1 (0.7) |  |
| CAM-S Score <sup>b</sup> | 8.2 (2.7) | 7.1 (3.0) | 9.3 (1.9) |  |
| Maximum CAM-S Score <sup>b</sup> | 12.9 (1.5) | 12.5 (1.6) | 13.3 (1.3) |  |
| Patients with CAM-S $\geq 7$ ever <sup>a</sup> | 62 (100) | 31 (100) | 31 (100) | |
| Patients with CAM-S $\geq 10$ ever <sup>a</sup> | 60 (97) | 29 (94) | 31 (100) | |
| Patients with CAM-S $\geq 12$ ever <sup>a</sup> | 53 (85) | 24 (77) | 29 (94) | |
| Days Delirium <sup>b</sup> | 12.7 (13.0) | 6.2 (4.3) | 19.3 (15.3) |  |
| Sum CAM-S <sup>b</sup> | 158.7 (137.0) | 89.6 (51.5) | 227.7 (160.2) |  |
| Opioids (Fentanyl Equivalents) (mg)<sup>c</sup> | 37.4 (78.9) | 32.7 (53.6) | *69.4 (82.0) | 346 (0.0296) |
| Benzodiazepines (Midazolam<br>Equivalents) (mg) <sup>c</sup> | 52.5 (813.3) | 8.0 (265.3) | **136.3 (1517.1) | 306.5 (0.0072) |
| Antipsychotics (g) <sup>c</sup> | 245 (723) | 130 (270) | ** 410 (879) | 281.0 (0.0025) |
| Propofol (g) <sup>c</sup> | 46 (53) | 36 (58) | * 54 (40) | 345.0 (0.0287) |
| Dexmedetomidine (mg) <sup>c</sup> | 6.3 (12.1) | 6.4 (11.4) | 6.2 (11.3) | 397.0 (0.1209) |
| Ketamine (mg) <sup>c</sup> | 0.0 (12291.8) | 0.0 (6138.5) | * 1971.6 (18993.5) | 359.0 (0.0323) |
| Days on Isoflurane <sup>b</sup> | 0.4 (1.1) | 0.4 (0.9) | 0.5 (1.3) | -0.6 (0.5611) |
| Days on Paralytics <sup>b</sup> | 6.8 (7.2) | 4.1 (3.0) | ** 9.5 (9.0) | -3.1 (0.0026) |
| **<u>Long-term Follow-up Data</u>** |  |  |  |  |
| Patients with Follow-up <sup>a</sup> | 40 (65) | 21 (68) | 19 (61) |  |
| TICS Score <sup>b</sup> | 28.9 (6.2) | 31.0 (5.0) | *26.3 (6.6) | 2.5 (0.018) |
| TICS Range | 12 – 39 | 22 – 39 | 12 – 36 |  |
| CES-D-10 Score <sup>b</sup> | 7.2 (5.1) | 6.7 (5.4) | 7.9 (4.6) | -0.8 (0.4566) |
| CES-D-10 Range | 0 – 22 | 0 – 22 | 0 – 14 |  |
| EQ-5D-3L Score <sup>b</sup> | 8.6 (2.5) | 7.7 (2.2) | **9.8 (2.3) | -2.9 (0.0062) |
| EQ-5D-3L Range | 5 – 14 | 5 – 11 | 6 – 14 |  |

<sup>a</sup> N (%)<sup>b</sup> Mean (Standard Deviation)<sup>c</sup> Median (IQR)
\* p-value < 0.05
\*\* p-value < 0.005

### Linear Regression

In **Table 3**, we show results using the TICS, CES-D-10, and EQ-5D-3L as outcome variables and ‘Days delirium’ as the exposure variable, while forcing the exposure variable to be included in step 1 of the stepwise variable selection. For TICS and CES-D-10, the forward selection included only ‘Days delirium’ into the regression model, and this single variable did not reach significance (TICS: F-test statistic 1.1, p=0.3; CES-D-10: F-test statistic 0.8, p=0.4). Pearson correlations were also non-significant between ‘Days delirium’ and TICS (r=-0.18, p=0.29); or between ‘Days delirium’ and CES-D-10 (r=0.22, p=0.20) (**Table S4**). For EQ-5D-3L, the regression model, consisting of the ‘Days delirium’ variable only, reached significance (F-test statistic 7.6, p=0.008), and adding hospital length of stay significantly improved the model’s fit (partial F-test statistic 9, p=0.005). The Pearson correlation between ‘Days delirium’ and EQ-5D-3L was moderate and significant (r=0.44, p=0.005).

**Table 3:** Linear Regression.

| TICS |  |  |  |  |  |  |  |  |  |
| --- | --- | --- | --- | --- | --- | --- | --- | --- | --- |
| Rank Included<br>Predictor Added | 0 – forced<br>Constant | 1 – forced<br>Days<br>Delirium | 2<br>Hospital<br>Length of<br>Stay | 3<br>Benzodiazep<br>ines and<br>Propofol | 4<br>Ketamine | 5<br>Opioids | 6<br>Sex | 7<br>BMI | 8<br>Age |
| R<sup>2</sup> | 0 | 0.03 | 0.14 | 0.19 | 0.24 | 0.25 | 0.26 | 0.26 | 0.26 |
| F-Statistic |  | 1.1 | 2.9 | 2.6 | 2.5 | 2.1 | 1.7 | 1.5 | 1.2 |
| F-test p-value |  | 0.3122 | 0.0705 | 0.0713 | 0.0586 | 0.0922 | 0.1515 | 0.2229 | 0.3164 |
| Log-Likelihood | -119.6 | -119 | -116.7 | -115.7 | -114.5 | -114.2 | -114.1 | -114 | -114 |
| Partial F-Statistic |  | 1.1 | 4.6 | 1.8 | 2.2 | 0.5 | 0.1 | 0.2 | 0 |
| Partial F-test p-value |  | 0.3122 | 0.0396 | 0.1867 | 0.1479 | 0.4927 | 0.7472 | 0.6886 | 0.8481 |
| Partial F-test p-value < 0.05 | N/A | FALSE | TRUE | FALSE | FALSE | FALSE | FALSE | FALSE | FALSE |
| CES-D-10 |  |  |  |  |  |  |  |  |  |
| Rank Included<br>Predictor Added | 0 – forced<br>Constant | 1 - forced<br>Days<br>Delirium | 2<br>BMI | 3<br>Benzodiazep<br>ines and<br>Propofol | 4<br>Hospital<br>Length of<br>Stay | 5<br>Age | 6<br>Sex | 7<br>Opioids | 8<br>Ketamine |
| R<sup>2</sup> | 0 | 0.02 | 0.23 | 0.31 | 0.36 | 0.39 | 0.41 | 0.41 | 0.42 |
| F-Statistic |  | 0.8 | 5.1 | 4.9 | 4.6 | 4.0 | 3.4 | 2.9 | 2.5 |
| F-test p-value |  | 0.3668 | 0.0117 | 0.0064 | 0.0050 | 0.0062 | 0.0110 | 0.0203 | 0.0348 |
| Log-Likelihood | -112 | -111.5 | -107.1 | -105.2 | -103.6 | -102.7 | -102.3 | -102.2 | -102.0 |
| Partial F-Statistic |  | 0.8 | 9.1 | 3.7 | 2.8 | 1.6 | 0.6 | 0.3 | 0.3 |
| Partial F-test p-value |  | 0.3668 | 0.0047 | 0.0637 | 0.1044 | 0.2152 | 0.4574 | 0.6067 | 0.6155 |
| Partial F-test p-value < 0.05 |  | FALSE | TRUE | FALSE | FALSE | FALSE | FALSE | FALSE | FALSE |
| EQ-5D-3L |  |  |  |  |  |  |  |  |  |
| Rank Included<br>Predictor Added | 0 – forced<br>Constant | 1 – forced<br>Days<br>Delirium | 2<br>Hospital<br>Length of<br>Stay | 3<br>Sex | 4<br>BMI | 5<br>Benzodiaze<br>pines and<br>Propofol | 6<br>Opioids | 7<br>Age | 8<br>Ketamine |
| R<sup>2</sup> | 0 | 0.17 | 0.34 | 0.39 | 0.42 | 0.43 | 0.44 | 0.45 | 0.45 |
| F-Statistic |  | 7.6 | 9.2 | 7.5 | 6.3 | 5 | 4.2 | 3.6 | 3 |
| F-test p-value |  | 0.0088 | 0.0006 | 0.0005 | 0.0007 | 0.0017 | 0.003 | 0.0064 | 0.0131 |
| Log-Likelihood | -90.2 | -86.5 | -82.2 | -80.5 | -79.4 | -79.3 | -78.8 | -78.7 | -78.7 |
| Partial F-Statistic |  | 7.6 | 9 | 3.1 | 2 | 0.3 | 0.8 | 0.1 | 0 |
| Partial F-test p-value |  | 0.0088 | 0.0049 | 0.0881 | 0.1661 | 0.615 | 0.3703 | 0.7411 | 0.8816 |
| Partial F-test p-value < 0.05 |  | TRUE | TRUE | FALSE | FALSE | FALSE | FALSE | FALSE | FALSE |

When the ‘Days delirium’ variable was not forced to be included in step 1, hospital length of stay was selected as the first variable for predicting TICS and EQ-5D-3L (TICS: F-test statistic 4.7, p=0.036; EQ-5D-3L: F-test statistic 16.4, p=0.0003). For CES-D-10, Body Mass Index (BMI) was selected as the first variable (F-test statistic 8.7, p=0.0056). For further details, see **Tables S5-S7**.

## Discussion

Our data show: 1) delirium early in the COVID-19 pandemic among acutely ill hospitalized patients was nearly universal, 2) patients received very large amounts of sedation and narcotics, and 3) delirium severity is associated with higher sedative doses, longer hospital and ICU stay, poorer quality of life and worse cognition at 3-month follow-up. Although we acknowledge that the association between delirium severity and poor outcome may not be direct and could be through illness severity, our findings document some of the largest quantities of sedation and narcotics administered to ICU populations and suggest the opportunity to improve sedation practices.

We observed a delirium prevalence of 98% (61 of the 62 patients) in our study, with 65% (40 patients) exhibiting the hyperactive subtype and 92% (57 patients) displaying the hypoactive subtype. The mean duration of delirium was 12.7 days. In comparison, previous studies have reported delirium prevalence rates ranging from 12.8% to 84.3% among ICU patients ^3,11,12,22-26^, with the hyperactive and hypoactive subtypes occurring in 12.6% to 86.6% and 13.4% to 87.4% of cases, respectively ^11,12,24^. By contrast, delirium prevalence among non-ICU patients has been reported at 22.8% to 33% ^27-29^. Historically, delirium prevalence in ICU patients has also been variable, with studies reporting 16.5%, 32.3%, 74%, and 84%, with mechanically ventilated patients composing 37%, 38.4%, 91%, and 100%, respectively ^10, 30-32^. A meta-analysis further estimated a pooled delirium prevalence of 31% among ICU patients, with hyperactive, hypoactive, and mixed subtypes occurring in 4%, 17%, and 10% of cases, respectively ^33^. This variability in reported prevalence can be attributed to differences in patient demographics, particularly those hospitalized during the COVID-19 pandemic, as well as variations in clinical settings, the criteria used to define delirium, and the screening tools employed across studies.

The use of sedatives is associated with delirium, especially sedative-hypnotics and anticholinergic agents ^34,35^. Heavy sedation, especially with benzodiazepines, is shown to increase the risk of delirium and coma in ICU patients ^36,37^, while minimizing sedation has led to improved survival and more delirium-free days ^38-40^. In our cohort, over 90% of patients received opioids, benzodiazepines, or propofol at least once, with higher cumulative doses compared to studies involving non-COVID-19 patients ^36,37^; and in line with other studies of COVID-19 patients ^11, 41^. One study reported that 64% and 70.9% of COVID-19 patients received benzodiazepines and propofol, respectively, for a median duration of 7 days ^11^.

The observed shift in practice from minimal sedation to deep sedation during the COVID-19 pandemic might be explained by various factors, including younger age and good health of many patients before the onset of COVID-19, high respiratory drive, shortage of trained ICU staff, and patients being more agitated on emergence from sedation ^42^. Also, individually tailored intermittent administration of certain drugs (opioids) may have been considered impractical in the face of the overwhelmed healthcare system during the early phase of the COVID-19 pandemic, and continuous sedation is practically favored, which increases the risk of side effects ^42^. In our cohort, 43 (64%) patients had ARDS due to COVID-19, which led to prolonged sedation and possibly drug tolerance, dependence, and other side effects, as well as severe delirium.

Symptoms of delirium that last for more than a few weeks or even months (persistent delirium) have been reported ^43^. Additionally, delirium is associated with an increased risk of mortality, institutionalization, and subsequent dementia ^43^. During 3-month follow-up, patients showed reductions in cognition, mood, and quality of life as measured by the TICS, CES-D-10, and EQ-5D-3L, respectively. These findings are consistent with a study on survivors of COVID-19-associated ARDS, which reported reduced quality of life, as measured by the EQ-5D-3L, in 67% of patients at a six-month follow-up ^44^. Such adverse outcomes after hospitalization may be related to the COVID-19 infection, critical illness, delirium, length of hospitalization, or, likely, a combination of these factors.

Prevention and management of delirium in critically ill COVID-19 patients, who are placed on enhanced respiratory isolation, presented unique challenges for clinicians. Techniques typically involve non-pharmacological interventions focused on reducing risk factors (e.g., benzodiazepine use), improving sleep quality, frequent reorientation, and increasing mobility ^45^. The ABCDEF bundle (Awakening and Breathing coordination, Choice of drugs, Delirium monitoring and management, Early exercise/mobility, and Family engagement) has been shown to improve survival and decrease delirium days in patients when followed ^40^. Due to the contagious nature of COVID-19 and new safety precautions put in place, many of these delirium treatment and prevention interventions became difficult to institute. Family visitation was typically not allowed, contributing to further social isolation and limiting the F element of the ABCDEF bundle to only interactions via online platforms. Additionally, sedation protocols were adjusted to deal with the large number of ARDS patients and prolonged mechanical ventilation. These factors likely put COVID-19 patients at higher risk for developing delirium during hospitalization.

Our study has several limitations. Importantly, a small sample size of 62 patients was able to be assessed for delirium. This study was conducted at a single center and included a population of 45% Hispanics, reducing cohort diversity/generalizability. All patients had at least one day with a CAM-S score ≥7. The lack of non-delirious patients limited the analysis of associations between delirium and long-term outcomes. When dichotomizing patients based on the median duration of delirium, we found significantly worse cognitive scores for the group with a higher delirium burden. Analogously, when splitting patients into tertiles, the top and middle tertiles had significantly worse cognitive scores than the bottom tertile. However, there was a low and insignificant correlation between the delirium exposure variables and cognitive scores, and a univariate linear regression model with delirium exposure (independent) and cognitive scores (dependent) did not fit the data significantly well. For quality-of-life, we similarly found significantly different scores when stratifying patients according to delirium exposure, but further observed significant correlation coefficients and univariate regression results. In stepwise variable selection, we found the length of hospital stay to be more strongly associated with cognitive and quality-of-life scores than delirium exposure. The non-significant associations between delirium and cognition, and length of hospitalization being a better predictor of cognition over delirium exposure, may be due to the small sample size and lack of a low delirium exposure control group. In contrast, prior research involving a larger cohort of 821 ICU patients found delirium duration to be independently associated with worse global cognition at three and twelve months post-discharge ^10^.

Overall, our study suggests that COVID-19 management guidelines should include delirium as a common presenting symptom, especially in elderly and ICU patients. There are several modifiable risk factors that may help to reduce delirium in COVID-19 patients and improve their quality-of-life post-discharge. One is the use of sedatives and their dosing and duration. The ability to minimize sedation in these critically ill patients is important for delirium management. Other risks may include a lack of family interaction due to visitor restrictions ^11^ and decreased re-orientation by clinical staff, due to policies that limit the amount of time spent in patient rooms as well as language barriers.

## Conclusion

Delirium and coma were very common in patients hospitalized for COVID-19 early in the pandemic. The total cumulative doses of sedating medications (including benzodiazepines, propofol, and paralytics) were associated with an increased severity of delirium. Additionally, patients experiencing more days of delirium often spent longer in the ICU and hospital, and on the ventilator. Finally, at 3-month follow-up, patients with more delirium days showed more impaired cognition and worse quality of life than those with fewer. To improve outcomes, we need to change the sedation practices of COVID-19 patients, adhere to the protocols that improve outcomes (ABCDEF bundle), and monitor for delirium.

## Acknowledgments

The authors wish to acknowledge and express their appreciation to the patients who were a part of this study, as well as the staff of the MGH ICUs and general care units who participated in their clinical care.

## Author contributions

JW and MBW conceived the study. AB, SQ, MA, SM, MM, and JW collected the clinical data. AB and WG analyzed the data. AB searched the scientific literature. AB and WG created figures. AB, WG, and RT drafted the initial manuscript. AB, JW, and MBW have verified the underlying data; had full access to all the data in the study, and take responsibility for the integrity of the data and the accuracy of the data analysis. All authors were involved in data interpretation and revising the manuscript for intellectual content.

## Declarations

### Conflicts of interest

MBW is a co-founder of Beacon Biosignals, which played no role in this work. All other authors report nothing to disclose.

### Funding

MBW was supported by the Glenn Foundation for Medical Research and American Federation for Aging Research (Breakthroughs in Gerontology Grant); American Academy of Sleep Medicine (AASM Foundation Strategic Research Award); Football Players Health Study (FPHS) at Harvard University; Department of Defense through a subcontract from Moberg ICU Solutions, Inc; and NIH (1R01NS102190, 1R01NS102574, 1R01NS107291, 1RF1AG064312, RF1NS120947, R01AG073410 R01HL161253, R01NS126282, R01AG073598), and NSF (2014431).

## Notes

Conflicts of interest: Dr. Westover is a co-founder, serves as a scientific advisor and consultant, and has a personal equity interest in Beacon Biosignals.

### Competing Interest Statement

Dr. Westover is a co-founder, serves as a scientific advisor and consultant, and has a personal equity interest in Beacon Biosignals.

### Author Declarations

According to the Partners Human Research Committee, it qualified for an exemption from formal oversight by the Mass General Brigham Institutional Review Board and met the criteria for a waiver of informed consent. The study was performed in compliance with the ethical principles outlined in the 1964 Declaration of Helsinki, its subsequent amendments, and comparable ethical standards.

## References

1. A Novel Coronavirus from Patients with Pneumonia in China, 2019 | NEJM [Internet]. [cited 2021 Mar 23] Available from: https://www.nejm.org/doi/10.1056/NEJMoa2001017?url_ver=Z39.88-2003&rfr_id=ori%3Arid%3Acrossref.org&rfr_dat=cr_pub++0 www.ncbi.nlm.nih.gov

2. CDC. COVID Data Tracker. Centers for Disease Control and Prevention. Published March 28, 2020. Accessed March 21, 2023. https://covid.cdc.gov/covid-data-tracker/#trends_totaldeaths_select_00

3. Helms J, Kremer S, Merdji H, et al.: Neurologic Features in Severe SARS-CoV-2 Infection [Internet]. N Engl J Med 2020; [cited 2021 Mar 5] Available from: https://www.ncbi.nlm.nih.gov/pmc/articles/PMC7179967/

4. American Psychiatric Association: Diagnostic and Statistical Manual of Mental Disorders [Internet]. Fifth Edition. American Psychiatric Association; 2013. [cited 2021 Mar 23] Available from: http://psychiatryonline.org/doi/book/10.1176/appi.books.9780890425596

5. Gibb K, Seeley A, Quinn T, et al.: The consistent burden in published estimates of delirium occurrence in medical inpatients over four decades: a systematic review and meta-analysis study. Age Ageing 2020; 49:352–360

6. Almeida ICT, Soares M, Bozza FA, et al.: The Impact of Acute Brain Dysfunction in the Outcomes of Mechanically Ventilated Cancer Patients [Internet]. PLoS One 2014; 9[cited 2021 Mar 5] Available from: https://www.ncbi.nlm.nih.gov/pmc/articles/PMC3899009/

7. Ely EW: Delirium as a Predictor of Mortality in Mechanically Ventilated Patients in the Intensive Care Unit. JAMA 2004; 291:1753

8. Hsieh SJ, Soto GJ, Hope AA, et al.: The Association between Acute Respiratory Distress Syndrome, Delirium, and In-Hospital Mortality in Intensive Care Unit Patients. Am J Respir Crit Care Med 2014; 191:71–78

9. Marengoni A, Zucchelli A, Grande G, et al.: The impact of delirium on outcomes for older adults hospitalised with COVID-19 [Internet]. Age Ageing 2020; [cited 2021 Mar 5] Available from: https://www.ncbi.nlm.nih.gov/pmc/articles/PMC7499475/

10. Pandharipande PP, Girard TD, Jackson JC, et al.: Long-Term Cognitive Impairment after Critical Illness. N Engl J Med 2013; 369:1306–1316

11. Prevalence and risk factors for delirium in critically ill patients with COVID-19 (COVID-D): a multicentre cohort study - The Lancet Respiratory Medicine [Internet]. [cited 2021 Mar 5] Available from: https://www.thelancet.com/journals/lanres/article/PIIS2213-2600(20)30552-X/fulltext

12. Helms J, Kremer S, Merdji H, et al.: Delirium and encephalopathy in severe COVID-19: a cohort analysis of ICU patients. Critical Care 2020; 24:491

13. Ely EW, Truman B, Shintani A, et al.: Monitoring Sedation Status Over Time in ICU Patients: Reliability and Validity of the Richmond Agitation-Sedation Scale (RASS). JAMA 2003; 289:2983

14. Ely EW, Inouye SK, Bernard GR, et al.: Delirium in Mechanically Ventilated Patients: Validity and Reliability of the Confusion Assessment Method for the Intensive Care Unit (CAM-ICU). JAMA 2001; 286:2703

15. Inouye SK, Kosar CM, Tommet D, et al.: The CAM-S: Development and Validation of a New Scoring System for Delirium Severity in 2 Cohorts. Ann Intern Med 2014; 160:526–533

16. Brandt J, Spencer M, Folstein M: The Telephone Interview for Cognitive Status. Cognitive and Behavioral Neurology 1988; 1:111–118

17. Irwin M, Artin KH, Oxman MN: Screening for Depression in the Older Adult: Criterion Validity of the 10-Item Center for Epidemiological Studies Depression Scale (CES-D). Arch Intern Med 1999; 159:1701

18. EuroQol Group: EuroQol--a new facility for the measurement of health-related quality of life. Health Policy 1990; 16:199–208

19. Graf C: The Lawton Instrumental Activities of Daily Living Scale. AJN The American Journal of Nursing 2008; 108:52–62

20. statsmodels.regression.linear_model.OLS — statsmodels [Internet]. [cited 2021 Mar 5] Available from: https://www.statsmodels.org/stable/generated/statsmodels.regression.linear_model.OLS.html

21. statsmodels.regression.linear_model.RegressionResults.compare_f_test — statsmodels [Internet]. [cited 2021 Mar 5] Available from: https://www.statsmodels.org/stable/generated/statsmodels.regression.linear_model.RegressionResults.compare_f_test.html

22. Fan S, Xiao M, Han F, et al.: Neurological Manifestations in Critically Ill Patients With COVID-19: A Retrospective Study [Internet]. Front Neurol 2020; 11[cited 2021 Apr 26] Available from: https://www.frontiersin.org/articles/10.3389/fneur.2020.00806/full

23. Jäckel M, Bemtgen X, Wengenmayer T, et al.: Is delirium a specific complication of viral acute respiratory distress syndrome? Critical Care 2020; 24:401

24. Khan SH, Lindroth H, Perkins AJ, et al.: Delirium Incidence, Duration, and Severity in Critically Ill Patients With Coronavirus Disease 2019 [Internet]. Crit Care Explor 2020; 2[cited 2021 Mar 5] Available from: https://www.ncbi.nlm.nih.gov/pmc/articles/PMC7690767/

25. Bernard-Valnet R, Favre E, Bernini A, et al. Delirium in Adults With COVID-19-related ARDS: Comparison With Other Etiologies [published online ahead of print, 2022 Aug 25]. Neurology. 2022;10.1212/WNL.0000000000201162. doi:10.1212/WNL.0000000000201162

26. Dias R, Caldas JP, Silva-Pinto A, Costa A, Sarmento A, Santos L. Delirium severity in critical patients with COVID-19 from an Infectious Disease Intensive Care Unit. Int J Infect Dis. 2022;118:109–115. doi:10.1016/j.ijid.2022.02.035

27. Garcez FB, Aliberti MJR, Poco PCE, et al.: Delirium and Adverse Outcomes in Hospitalized Patients with COVID-19. Journal of the American Geriatrics Society 2020; 68:2440–2446

28. Wilke V, Sulyok M, Stefanou MI, et al. Delirium in hospitalized COVID-19 patients: Predictors and implications for patient outcome. PLoS One. 2022;17(12):e0278214. Published 2022 Dec 22. doi:10.1371/journal.pone.0278214

29. Callea A, Conti G, Fossati B, et al. Delirium in hospitalized patients with COVID-19 pneumonia: a prospective, cross-sectional, cohort study. Intern Emerg Med. 2022;17(5):1445–1452. doi:10.1007/s11739-022-02934-w

30. Salluh JI, Soares M, Teles JM, et al.: Delirium epidemiology in critical care (DECCA): an international study. Crit Care 2010; 14:R210

31. Khan SH, Lindroth H, Hendrie K, et al.: Time trends of delirium rates in the intensive care unit. Heart Lung 2020; 49:572–577

32. Brummel NE, Jackson JC, Pandharipande PP, et al.: Delirium in the Intensive Care Unit and Subsequent Long-term Disability Among Survivors of Mechanical Ventilation. Crit Care Med 2014; 42:369–377

33. Krewulak KD, Stelfox HT, Leigh JP, et al.: Incidence and Prevalence of Delirium Subtypes in an Adult ICU: A Systematic Review and Meta-Analysis*. Critical Care Medicine 2018; 46:2029–2035

34. Pandharipande PP, Shintani A, Peterson J, et al.: SEDATIVE AND ANALGESIC MEDICATIONS ARE INDEPENDENT RISK FACTORS IN ICU PATIENTS FOR TRANSITIONING INTO DELIRIUM: 75. Critical Care Medicine 2004; 32:A19

35. Han L, McCusker J, Cole M, et al.: Use of medications with anticholinergic effect predicts clinical severity of delirium symptoms in older medical inpatients. Arch Intern Med 2001; 161:1099–1105

36. Campbell NL, Perkins AJ, Khan BA, et al.: Deprescribing in the Pharmacologic Management of Delirium: A Randomized Trial in the Intensive Care Unit. J Am Geriatr Soc 2019; 67:695–702

37. Pharmacoeconomic Modeling of Lorazepam, Midazolam, and Propofol for Continuous Sedation in Critically Ill Patients - MacLaren - 2005 - Pharmacotherapy: The Journal of Human Pharmacology and Drug Therapy - Wiley Online Library [Internet]. [cited 2021 Mar 8] Available from: https://accpjournals.onlinelibrary.wiley.com/doi/abs/10.1592/phco.2005.25.10.1319

38. Pun BT, Balas MC, Barnes-Daly MA, et al.: Caring for Critically Ill Patients with the ABCDEF Bundle: Results of the ICU Liberation Collaborative in Over 15,000 Adults. Crit Care Med 2019; 47:3–14

39. Barnes-Daly MA, Pun BT, Harmon LA, et al.: Improving Health Care for Critically Ill Patients Using an Evidence-Based Collaborative Approach to ABCDEF Bundle Dissemination and Implementation. Worldviews Evid Based Nurs 2018; 15:206–216

40. Barnes-Daly MA, Phillips G, Ely EW: Improving Hospital Survival and Reducing Brain Dysfunction at Seven California Community Hospitals: Implementing PAD Guidelines Via the ABCDEF Bundle in 6,064 Patients. Crit Care Med 2017; 45:171–178

41. Benghanem S, Cariou A, Diehl JL, et al. Early Clinical and Electrophysiological Brain Dysfunction Is Associated With ICU Outcomes in COVID-19 Critically Ill Patients With Acute Respiratory Distress Syndrome: A Prospective Bicentric Observational Study. Crit Care Med. 2022;50(7):1103–1115. doi:10.1097/CCM.0000000000005491

42. Hanidziar D, Bittner EA: Sedation of Mechanically Ventilated COVID-19 Patients: Challenges and Special Considerations [Internet]. Anesth Analg 2020; [cited 2021 Mar 5] Available from: https://www.ncbi.nlm.nih.gov/pmc/articles/PMC7179055/

43. Wilson JE, Mart MF, Cunningham C, et al.: Delirium. Nature Reviews Disease Primers 2020; 6:1– 26

44. Jacobs LG, Paleoudis EG, Bari DL-D, et al.: Persistence of symptoms and quality of life at 35 days after hospitalization for COVID-19 infection. PLOS ONE 2020; 15:e0243882

45. Clinical Practice Guidelines for the Prevention and Manageme…L: Critical Care Medicine [Internet]. [cited 2021 Mar 23] Available from: https://journals.lww.com/ccmjournal/Fulltext/2018/09000/Clinical_Practice_Guidelines_for_the_Pr evention.29.aspx

46. Charlson ME, Pompei P, Ales KL, MacKenzie CR. A new method of classifying prognostic comorbidity in longitudinal studies: development and validation. J Chronic Dis. 1987;40(5):373–83

47. Ferreira FL, Bota DP, Bross A, Mélot C, Vincent JL. Serial evaluation of the SOFA score to predict outcome in critically ill patients. JAMA. 2001 Oct 10;286(14):1754–8.

