## Supplement File for "Delirium and High Sedation Levels are Common in Critically Ill Patients with COVID-19 and Associated with Poor Long-Term Outcomes"

Corresponding Author:

M. Brandon Westover, MD, PhD

**Table of contents**

**Supplemental Methods:**

- Delirium Prevalence
- Medications
- Linear Regression

**Supplemental Results:**

**Supplemental Figures**

**Figure S1:** Flow diagram of subject enrollment

**Figure S2:** Swimmer plots indicating the CAM-S scores for each study patient

**Figure S3:** Swimmer plots indicating the RASS scores for each study patient

**Supplemental Tables**

**Table S1:** Exposure Strata – ‘Days Delirium’, comparing the moderate and severe groups to the mild group

**Table S2**: Exposure Strata – ‘Days Delirium’

**Table S3:** Exposure Strata – ‘Sum CAM-S’

**Table S4:** Pearson correlation coefficients between delirium and outcome variables

**Table S5:** Linear Regression – Cognition (TICS) Not Forced

**Table S6:** Linear Regression – Depression (CES-D-10) Not Forced

**Table S7**: Linear Regression – Quality of Life (EQ-5D-3L) Not Forced

**Supplemental References**

### **Methods**

#### Delirium Prevalence – ‘Days delirium fraction’ is determined by taking the number of days delirious divided by the total number of days assessed. ‘Sum CAM-S fraction’ is the sum of the CAM-S scores divided by the total number of days assessed. ‘Sum RASS’ includes the sum of all RASS scores, including both positive and negative RASS scores. ‘Sum RASS fraction’ is the sum of all RASS scores divided by the total number of days with RASS score data (admission to discharge).

#### Medications – Medication equivalent conversions were done as follows: opioids were converted to fentanyl equivalents **^1^**, and the following sedatives were converted to midazolam equivalents as shown: 0.5 mg lorazepam = 1 mg midazolam = 10 mg chlordiazepoxide = 0.125 mg clonazepam = 0.25 mg alprazolam = 1.25 mg diazepam **^2,3^**.

#### Linear Regression – We performed a stepwise multivariable regression procedure using forward variable selection with the partial F-test **^4,5^**. Long-term follow-up outcomes were treated as dependent variables, and delirium prevalence, the main exposure of interest, and covariates as independent variables. A Box Hox transformation was applied to medication variables. All variables were scaled to a mean = 0 and standard deviation = 1. Ordinary Least Squares (OLS) method was used to fit each individual multivariate regression model. We ran the stepwise regression procedure twice, once as is and once where we forced the exposure variable to be included first after the constant variable. The model runs as follows:

1. Train the OLS with only a constant variable included (baseline model).
2. Train a model with the variable already included (constant variable only for step 1) and one more variable. Do this for all of the variables.
3. For each model, run an F-test to compare the two models:
   i. Ho, Null Hypothesis: The model including the new variable does NOT explain more variance in the dependent variable than the ‘old’ model.
   ii. H1, Alternate Hypothesis: The new model including the new variable does explain more variance in the dependent variable than the ‘old’ model.
   iii. The F-test provides the F-statistic and p-value as test results.
4. Select the variable that achieved the highest F-statistic in step 3 and report the F-test result (especially if the p-value was <0.05, i.e., if there was a significant improvement when including the variable). Report the results of the model including the newly selected variable (see Tables S4-7).
5. Repeat Step 2, starting with the model including the newly selected variable, until all variables are included in the model.

We run the model stepwise regression procedure once where we force the exposure variable to be included first after the constant variable, and then continue running the procedure in the same manner described above for all other steps.

**Figure S1:** Flow diagram of subject enrollment

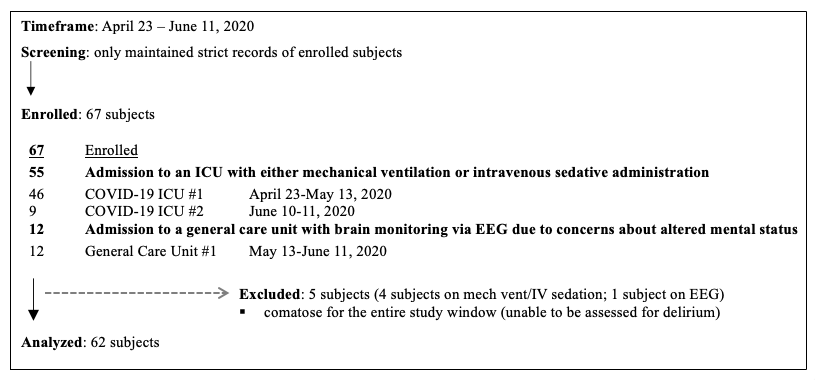

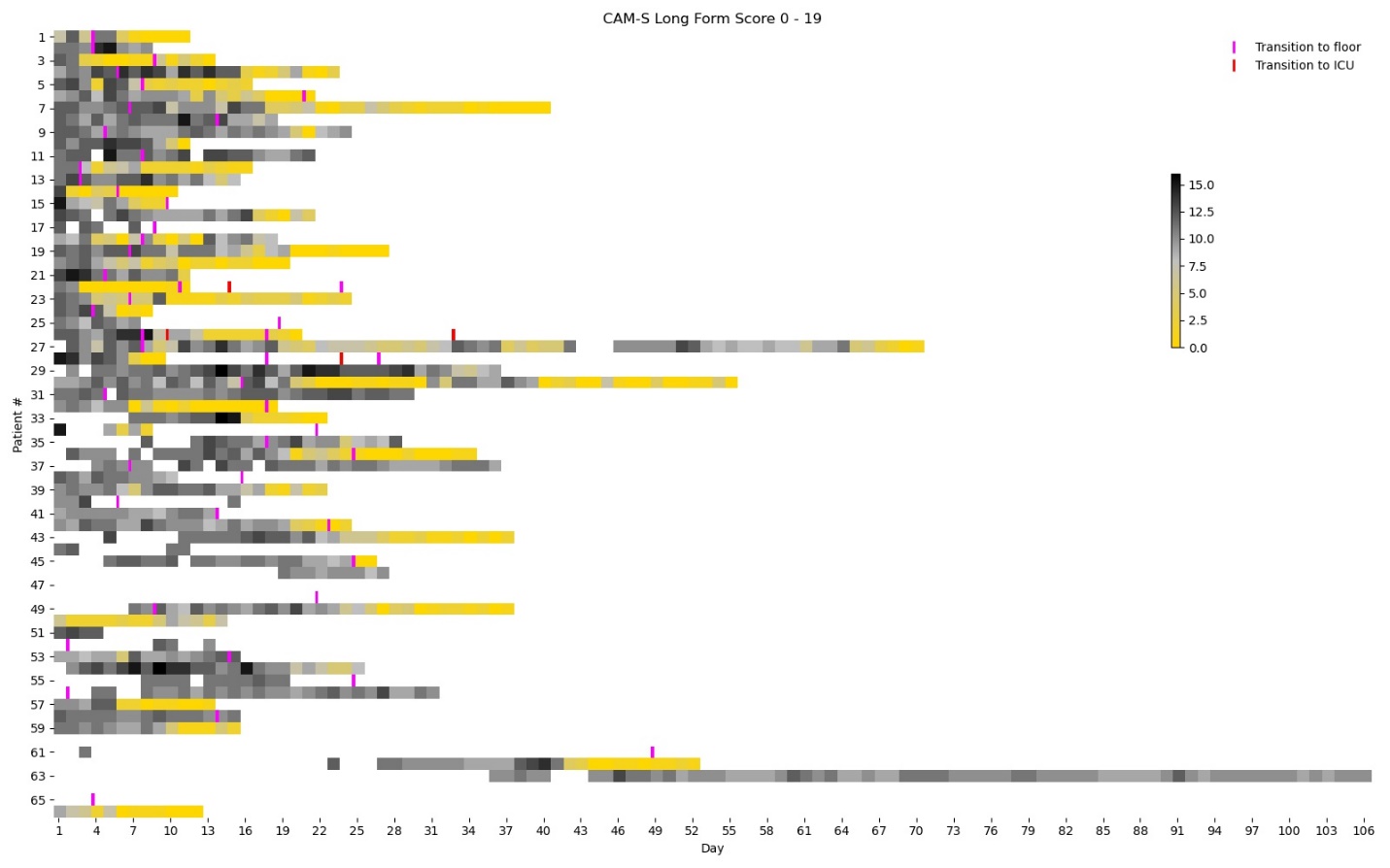

**Figure S2:** Swimmer plots indicating the CAM-S scores for each study patient. Each bin represents one day. Transitions from the ICU to the floor, and from the floor back to the ICU are also shown.

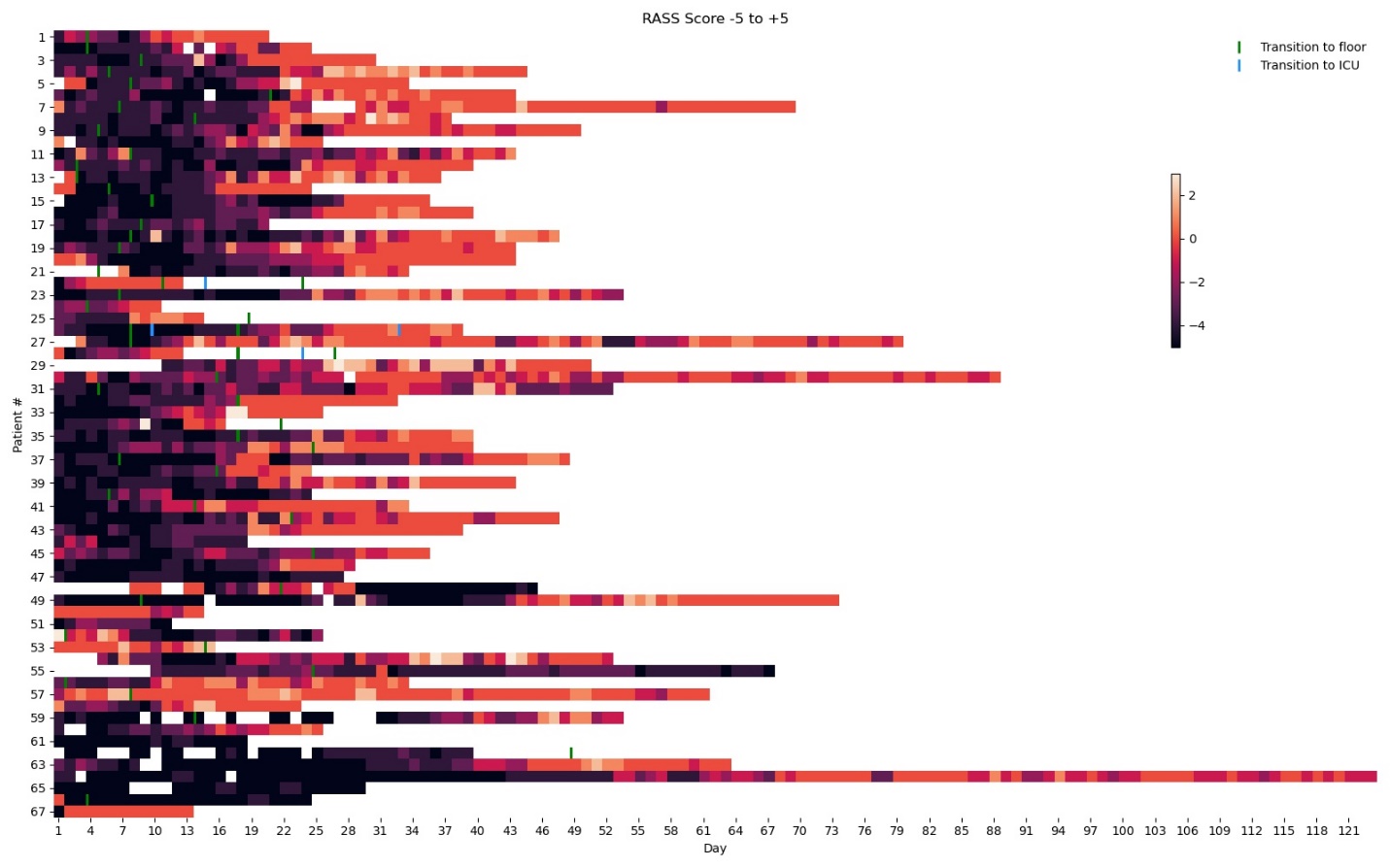

**Figure S3:** Swimmer plots indicating the RASS scores for each study patient. Each bin represents one day. Transitions from the ICU to the floor, and from the floor back to the ICU are also shown. RASS score data was obtained from admission to discharge, so day 1 is the day of hospital admission.

**Table S1:** Exposure Strata – ‘Days Delirium’, comparing the moderate and severe groups to the mild group

|  | All Patients | Days Delirium or Coma ≤ 20  (Mild) | 20 < Days Delirium or Coma ≤ 29.9  (Moderate) | Days Delirium or Coma > 29.9 (Severe) | Compare moderate to mild t-statistic (p-value), U-statistic for medications | Compare severe to mild t-statistic (p-value), U-statistic for medications |
| --- | --- | --- | --- | --- | --- | --- |
| **Hospitalization Data** |  |  |  |  |  |  |
| N Patients | 62 | 20 | 21 | 21 |  |  |
| Sex  Male ^a^  Female ^a^  Age (years) ^b^ | 40 (60)  27 (40)  59 (13) | 10 (50)  10 (50)  56 (15) | 13 (60)  8 (40)  58 (12) | 15 (70)  6 (30)  61 (11) | -0.8 (0.4552)  -0.5 (0.6) | -1.1 (0.2905)  (-1.2, 0.2) |
| BMI (kg/m^2^) ^b^ | 30.7 (7.0) | 32.0 (6.6) | 31.2 (8.2) | 28.8 (5.9) | 0.3 (0.7546) | 1.6 (0.1171) |
| ICU Length of Stay (days) ^b^ | 22.4 (13.0) | 13.2 (6.9) | **24.8 (8.1) | **29.6 (16.2) | -4.6 (<0.0001) | -4.2 (0.0002) |
| Hospital Length of Stay (days) ^b^ | 38.0 (20.6) | 21.2 (7.9) | **40.8 (15.0) | **51.0 (23.5) | -5.2 (<0.0001) | -5.4 (<0.0001) |
| Days Intubated ^b^ | 18.9 (11.4) | 12.4 (6.2) | *23.7 (15.8) | **20.0 (2.3) | -2.7 (0.011) | -4.4 (0.0002) |
| Days Intubated or Tracheostomy ^b^ | 22.9 (13.5) | 12.7 (6.7) | **27.7 (15.8) | **27.9 (9.0) | -3.5 (0.0012) | -5.3 (<0.0001) |
| CCI Score ^b^ | 1.2 (1.5) | 1.6 (1.8) | 1.1 (0.9) | 1.0 (1.8) | 1.1 (0.2621) | 1.1 (0.2896) |
| Mortality ^a^ | 10 (16) | 2 (10) | 3 (14) | 5 (20) | -0.4 (0.68) | -1.2 (0.25) |
| SOFA Score ^b^ | 5.1 (4.0) | 5.1 (4.0) | 3.8 (3.7) | 6.5 (4.1) | 1.1 (0.2676) | -1.1 (0.2832) |
| Patients with CAM-ICU ≥ 3 ever ^a^ | 61 (98) | 19 (95) | 21 (100) | 21 (100) |  |  |
| CAM-ICU Score ^b^ | 2.7 (1.0) | 2.1 (1.1) | 2.7 (0.9) | 3.4 (0.6) |  |  |
| CAM-S Score ^b^ | 8.2 (2.7) | 6.5 (3.2) | 8.2 (2.3) | 9.9 (1.5) |  |  |
| Maximum CAM-S Score ^b^ | 12.9 (1.5) | 12.4 (1.8) | 13.0 (1.5) | 13.3 (1.1) |  |  |
| Patients with CAM-S ≥ 7 ever ^a^ | 62 (100) | 20 (100) | 21 (100) | 21 (100) |  |  |
| Patients with CAM-S ≥ 10 ever ^a^ | 60 (97) | 18 (90) | 21 (100) | 21 (100) |  |  |
| Patients with CAM-S ≥ 12 ever ^a^ | 53 (85) | 15 (75) | 17 (81) | 21 (100) |  |  |
| Days Delirium ^b^ | 12.7 (13.0) | 4.4 (3.1) | 10.0 (5.5) | 23.3 (16.7) |  |  |
| Days Delirium Fraction ^b^ | 0.5 (0.3) | 0.4 (0.3) | 0.5 (0.3) | 0.7 (0.2) |  |  |
| Sum CAM-S ^b^ | 158.7 (137.0) | 66.0 (31.8) | 142.7 (74.3) | 262.8 (174.8) |  |  |
| Sum CAM-S Fraction ^b^ | 6.9 (2.8) | 5.5 (2.7) | 6.8 (2.8) | 8.2 (2.3) |  |  |
| Sum RASS ^b^ | -59.6 (44.7) | -29.2 (22.0) | -65.8 (27.5) | -82.5 (58.0) |  |  |
| Sum RASS Fraction ^b^ | -1.6 (0.9) | -1.3 (0.9) | -1.8 (0.9) | -1.6 (0.7) |  |  |
| Opioids (Fentanyl Equivalents) (mg)^c^ | 37.4 (78.9) | 27.6 (36.2) | **69.9 (72.1) | 43.7 (78.5) | 112.0 (0.0055) | 147.0 (0.0515) |
| Benzodiazepines (Midazolam Equivalents) (mg) ^c^ | 52.5 (813.3) | 8.0 (96.1) | *149.5 (959.1) | *103.2 (989.9) | 143.0 (0.0409) | 129.5 (0.0183) |
| Antipsychotics (g) ^c^ | 245 (722) | 95 (284) | **320 (750) | **410 (850) | 113.5 (0.0061) | 113.0 (0.0059) |
| Propofol (g) ^c^ | 46 (53) | 13 (41) | **51 (41) | **57 (40) | 103.0 (0.0027) | 115.0 (0.0068) |
| Dexmedetomidine (mg) ^c^ | 6.3 (12.1) | 6.6 (15.6) | 5.5 (8.9) | 6.2 (12.5) | 198.0 (0.3818) | 185.0 (0.2609) |
| Ketamine (mg) ^c^ | 0.0 (12291.8) | 0.0 (3345.0) | *1574.4 (19100.2) | *1971.6 (17586.5) | 145.0 (0.0319) | 149.0 (0.0388) |
| Days on Isoflurane ^b^ | 0.4 (1.1) | 0.2 (0.6) | 0.7 (1.5) | 0.3 (1.0) | -1.3 (0.1972) | -0.3 (0.7476) |
| Days on Paralytics ^b^ | 6.8 (7.2) | 2.7 (2.2) | ** 8.2 (6.1) | **9.3 (9.5) | -3.8 (0.0005) | -3.0 (0.0043) |
| **Long-term Follow-up Data** |  |  |  |  |  |  |
| Patients with Follow-up ^a^ | 40 (65) | 14 (70) | 13 (62) | 13 (62) |  |  |
| TICS Score ^b^ | 28.9 (6.2) | 30.3 (4.9) | 27.8 (8.1) | 28.5 (5.3) | 1.0 (0.341) | 0.9 (0.3811) |
| TICS Range | 12 – 39 | 22 – 38 | 12 – 39 | 16 - 36 |  |  |
| CES-D-10 Score ^b^ | 7.2 (5.1) | 6.7 (6.1) | 6.7 (4.2) | 8.5 (4.6) | 0.0 (0.9821) | -0.8 (0.4422) |
| CES-D-10 Range | 0 – 22 | 0 – 22 | 1 – 14 | 0 – 13 |  |  |
| EQ-5D-3L Score ^b^ | 8.6 (2.5) | 7.1 (2.2) | 8.6 (2.5) | **10.3 (1.8) | -1.6 (0.1268) | -4.1 (0.0003) |
| EQ-5D-3L Range | 5 – 14 | 5 – 11 | 6 – 14 | 7 – 13 |  |  |

^a^ N (%)

^b^ Mean (Standard Deviation)

^c^ Median (IQR)

*	p-value < 0.05

** p-value < 0.005

**Table S2**: Exposure Strata – ‘Days Delirium’

|  | All Patients | Days Delirium ≤ 10 | Days Delirium > 10 | t-statistic (p-value)  U-statistic for medications |
| --- | --- | --- | --- | --- |
| **Hospitalization Data** |  |  |  |  |
| N Patients | 62 | 30 | 32 |  |
| Sex  Male ^a^  Female ^a^ | 40 (60)  27 (40) | 18 (60)  12 (40) | 19 (60)  13 (40) | -0.5 (0.6464) |
| Age (years) ^b^ | 59 (13) | 55 (13) | 61 (12) | -1.9 (0.057) |
| BMI (kg/m^2^) ^b^ | 30.7 (7.0) | 31.7 (7.1) | 29.7 (6.9) | 1.1 (0.2764) |
| ICU Length of Stay (days) ^b^ | 22.4 (13.0) | 18.0 (9.1) | **26.5 (14.9) | -2.7 (0.0094) |
| Hospital Length of Stay (days) ^b^ | 38.0 (20.6) | 24.9 (10.8) | **50.2 (20.2) | -6.1 (<0.0001) |
| Days Intubated ^b^ | 18.9 (11.4) | 15.2 (6.6) | *22.9 (14.0) | -2.5 (0.0164) |
| Days Intubated or Tracheostomy ^b^ | 22.9 (13.5) | 17.0 (9.1) | **29.0 (14.6) | -3.5 (0.0011) |
| CCI Score ^b^ | 1.2 (1.5) | 1.4 (1.6) | 1.0 (1.5) | 1.0 (0.3084) |
| Mortality ^a^ | 10 (16) | 7 (23) | 3 (9) | 1.5 (0.14) |
| SOFA Score ^b^ | 5.1 (4.0) | 5.3 (3.8) | 4.9 (4.3) | 0.4 (0.7257) |
| Patients with CAM-ICU ≥ 3 ever ^a^ | 61 (98) | 29 (97) | 32 (100) |  |
| CAM-ICU Score ^b^ | 2.7 (1.0) | 2.4 (1.2) | 3.0 (0.7) |  |
| CAM-S Score ^b^ | 8.2 (2.7) | 7.3 (3.2) | 9.1 (1.8) |  |
| Maximum CAM-S Score ^b^ | 12.9 (1.5) | 12.4 (1.7) | 13.3 (1.2) |  |
| Patients with CAM-S ≥ 7 ever ^a^ | 62 (100) | 30 (100) | 32 (100) |  |
| Patients with CAM-S ≥ 10 ever ^a^ | 60 (97) | 28 (93) | 32 (100) |  |
| Patients with CAM-S ≥ 12 ever ^a^ | 53 (85) | 22 (73) | 31 (97) |  |
| Days Delirium ^b^ | 12.7 (13.0) | 4.3 (2.6) | 20.6 (13.9) |  |
| Days Delirium Fraction ^b^ | 0.5 (0.3) | 0.4 (0.3) | 0.7 (0.2) |  |
| Sum CAM-S ^b^ | 158.7 (137.0) | 66.0 (30.2) | 245.6 (141.3) |  |
| Sum CAM-S Fraction ^b^ | 6.9 (2.8) | 5.5 (2.7) | 8.2 (2.2) |  |
| Sum RASS ^b^ | -59.6 (44.7) | -48.2 (33.7) | -70.4 (51.2) |  |
| Sum RASS Fraction ^b^ | -1.6 (0.9) | -1.7 (1.0) | -1.4 (0.7) |  |
| Opioids (Fentanyl Equivalents) (mg)^c^ | 37.4 (78.9) | 38.2 (75.1) | 33.4 (75.5) | 458.0 (0.381) |
| Benzodiazepines (Midazolam Equivalents) (mg) ^c^ | 52.5 (813.3) | 28.8 (815.0) | 100.9 (735.3) | 388.5 (0.0994) |
| Antipsychotics (g) ^c^ | 245 (723) | 120 (282) | **400 (837) | 289.5 (0.0037) |
| Propofol (g) ^c^ | 46 (53) | 36 (54) | *56 (38) | 357.0 (0.0422) |
| Dexmedetomidine (mg) ^c^ | 6.3 (12.1) | 6.6 (13.3) | 5.9 (11.5) | 460.0 (0.3916) |
| Ketamine (mg) ^c^ | 0.0 (12291.8) | 0.0 (10167.0) | 38.6 (12634.0) | 456.0 (0.3597) |
| Days on Isoflurane ^b^ | 0.4 (1.1) | 0.5 (1.0) | 0.4 (1.2) | 0.2 (0.8280) |
| Days on Paralytics ^b^ | 6.8 (7.2) | 5.4 (6.0) | 8.1 (8.0) | -1.5 (0.1367) |
| **Long-term Follow-up Data** |  |  |  |  |
| Patients with Follow-up ^a^ | 40 (65) | 19 (63) | 21 (66) |  |
| TICS Score ^b^ | 28.9 (6.2) | 30.3 (5.9) | 27.5 (6.4) | 1.4 (0.1646) |
| TICS Range | 12 – 39 | 17 – 39 | 12 – 37 |  |
| CES-D-10 Score ^b^ | 7.2 (5.1) | 6.5 (5.7) | 7.9 (4.4) | -0.8 (0.4093) |
| CES-D-10 Range | 0 – 22 | 0 – 22 | 0 – 14 |  |
| EQ-5D-3L Score ^b^ | 8.6 (2.5) | 7.2 (2.1) | **9.9 (2.2) | -3.9 (0.0004) |
| EQ-5D-3L Range | 5 – 14 | 5 – 11 | 6 – 14 |  |

^a^ N (%)

^b^ Mean (Standard Deviation)

^c^ Median (IQR)

*	p-value < 0.05

** p-value < 0.005

**Table S3:** Exposure Strata – ‘Sum CAM-S’

|  | All Patients | Sum CAM-S ≤ 121 | Sum CAM-S >121 | t-statistic (p-value)  U-statistic for medications |
| --- | --- | --- | --- | --- |
| **Hospitalization Data** |  |  |  |  |
| N Patients | 62 | 31 | 31 |  |
| Sex  Male ^a^  Female ^a^ | 40 (60)  27 (40) | 15.5 (50)  15.5 (50) | 19 (60)  12 (40) | -0.8 (0.4456) |
| Age (years) ^b^ | 59 (13) | 55 (13) | 62 (12) | -1.9 (0.0586) |
| BMI (kg/m^2^) ^b^ | 30.7 (7.0) | 31.2 (7.4) | 30.1 (6.7) | 0.7 (0.5150) |
| ICU Length of Stay (days) ^b^ | 22.4 (13.0) | 18.1 (8.9) | **26.7 (15.1) | -2.7 (0.0087) |
| Hospital Length of Stay (days) ^b^ | 38.0 (20.6) | 25.1 (10.7) | **50.8 (20.3) | -6.2 (<0.0001) |
| Days Intubated ^b^ | 18.9 (11.4) | 15.3 (6.5) | *23.0 (14.3) | -2.5 (0.0175) |
| Days Intubated or Tracheostomy ^b^ | 22.9 (13.5) | 17.3 (9.0) | **29.2 (14.9) | -3.4 (0.0013) |
| CCI Score ^b^ | 1.2 (1.5) | 1.4 (1.6) | 1.1 (1.5) | 0.8 (0.4143) |
| Mortality ^a^ | 10 (16) | 7 (23) | 3 (10) | 1.4 (0.1727) |
| SOFA Score ^b^ | 5.1 (4.0) | 5.1 (3.8) | 5.1 (4.3) | 0.0 (0.9751) |
| Patients with CAM-ICU ≥ 3 ever ^a^ | 61 (98) | 30 (97) | 31 (100) |  |
| CAM-ICU Score ^b^ | 2.7 (1.0) | 2.4 (1.2) | 3.0 (0.7) |  |
| CAM-S Score ^b^ | 8.2 (2.7) | 7.4 (3.3) | 9.0 (1.8) |  |
| Maximum CAM-S Score ^b^ | 12.9 (1.5) | 12.5 (1.7) | 13.3 (1.2) |  |
| Patients with CAM-S ≥ 7 ever ^a^ | 62 (100) | 31 (100) | 31 (100) |  |
| Patients with CAM-S ≥ 10 ever ^a^ | 60 (97) | 29 (94) | 31 (100) |  |
| Patients with CAM-S ≥ 12 ever ^a^ | 53 (85) | 23 (74) | 30 (97) |  |
| Days Delirium ^b^ | 12.7 (13.0) | 4.5 (2.8) | 20.9 (13.9) |  |
| Days Delirium Fraction ^b^ | 0.5 (0.3) | 0.4 (0.3) | 0.7 (0.2) |  |
| Sum CAM-S ^b^ | 158.7 (137.0) | 67.7 (31.2) | 249.6 (141.7) |  |
| Sum CAM-S Fraction ^b^ | 6.9 (2.8) | 5.7 (2.8) | 8.1 (2.1) |  |
| Sum RASS ^b^ | -59.6 (44.7) | -48.6 (33.3) | -70.6 (52.0) |  |
| Sum RASS Fraction ^b^ | -1.6 (0.9) | -1.7 (1.0) | -1.4 (0.7) |  |
| Opioids (Fentanyl Equivalents) (mg)^c^ | 37.4 (78.9) | 38.8 (80.7) | 32.9 (71.3) | 476.0 (0.4775) |
| Benzodiazepines (Midazolam Equivalents) (mg) ^c^ | 52.5 (813.3) | 47.6 (793.6) | 98.6 (798.2) | 398.5 (0.1251) |
| Antipsychotics (g) ^c^ | 245 (723) | 130 (287) | **390 (885) | 302.5 (0.0062) |
| Propofol (g) ^c^ | 46 (53) | 36 (51) | *57 (39) | 352.0 (0.0358) |
| Dexmedetomidine (mg) ^c^ | 6.3 (12.1) | 6.4 (13.1) | 6.2 (11.5) | 474.0 (0.4663) |
| Ketamine (mg) ^c^ | 0.0 (12291.8) | 0.0 (9751.1) | 77.3 (12785.9) | 442.0 (0.2808) |
| Days on Isoflurane ^b^ | 0.4 (1.1) | 0.5 (1.0) | 0.4 (1.2) | 0.1 (0.9076) |
| Days on Paralytics ^b^ | 6.8 (7.2) | 5.4 (5.9) | 8.3 (8.1) | -1.6 (0.1122) |
| **Long-term Follow-up Data** |  |  |  |  |
| Patients with Follow-up ^a^ | 40 (65) | 20 (65) | 20 (65) |  |
| TICS Score ^b^ | 28.9 (6.2) | 30.0 (5.9) | 27.7 (6.5) | 1.1 (0.259) |
| TICS Range | 12 – 39 | 17 – 39 | 12 – 37 |  |
| CES-D-10 Score ^b^ | 7.2 (5.1) | 6.4 (5.6) | 8.1 (4.5) | -1.0 (0.3327) |
| CES-D-10 Range | 0 – 22 | 0 – 22 | 0 – 14 |  |
| EQ-5D-3L Score ^b^ | 8.6 (2.5) | 7.4 (2.1) | **9.8 (2.2) | -3.6 (0.001) |
| EQ-5D-3L Range | 5 – 14 | 5 – 11 | 6 – 14 |  |

^a^ N (%)

^b^ Mean (Standard Deviation)

^c^ Median (IQR)

*	p-value < 0.05

** p-value < 0.005

**Table S4:** Pearson correlation coefficients between delirium and outcome variables

|  | TICs | CES-D-10 |  | EQ-5D-3L |
| --- | --- | --- | --- | --- |
| N Patients | 37 | 37 |  | 39 |
| Delirium Days | -0.18 (p=0.29) | 0.22 (p=0.20) |  | 0.44 (p=0.005) |
| Sum CAM-S | -0.22 (p=0.19) | 0.26 (p=0.12) |  | 0.46 (p=0.003) |

**Table S5:** Linear Regression – Cognition (TICS) Not Forced

| TICS | | | | | | | | | |
| --- | --- | --- | --- | --- | --- | --- | --- | --- | --- |
| Rank Included | 0 – forced | 1 | 2 | 3 | 4 | 5 | 6 | 7 | 8 |
| Predictor Added | Constant | Hospital Length of Stay | Benzodiazepines and Propofol | Ketamine | Days Delirium | Opioids | Sex | BMI | Age |
| R^2^ | 0 | 0.12 | 0.18 | 0.22 | 0.24 | 0.25 | 0.26 | 0.26 | 0.26 |
| F-Statistic |  | 4.7 | 3.8 | 3.1 | 2.5 | 2.1 | 1.7 | 1.5 | 1.2 |
| F-test p-value |  | 0.0364 | 0.0318 | 0.0414 | 0.0586 | 0.0922 | 0.1515 | 0.2229 | 0.3164 |
| Log-Likelihood | -119.6 | -117.2 | -115.8 | -115 | -114.5 | -114.2 | -114.1 | -114 | -114 |
| Partial F-Statistic |  | 4.7 | 2.7 | 1.4 | 1 | 0.5 | 0.1 | 0.2 | 0 |
| Partial F-test p-value |  | 0.0364 | 0.1106 | 0.2372 | 0.3290 | 0.4927 | 0.7472 | 0.6886 | 0.8481 |
| Partial F-test p-value < 0.05 | N/A | TRUE | FALSE | FALSE | FALSE | FALSE | FALSE | FALSE | FALSE |

**Table S6:** Linear Regression – Depression (CES-D-10) Not Forced

| CES-D-10 | | | | | | | | | |
| --- | --- | --- | --- | --- | --- | --- | --- | --- | --- |
| Rank Included | 0 – forced | 1 - | 2 | 3 | 4 | 5 | 6 | 7 | 8 |
| Predictor Added | Constant | BMI | Benzodiazepines and Propofol | Ketamine | Age | Opioids | Days Delirium | Hospital Length of Stay | Sex |
| R^2^ | 0 | 0.20 | 0.31 | 0.34 | 0.36 | 0.37 | 0.38 | 0.41 | 0.42 |
| F-Statistic |  | 8.7 | 7.5 | 5.6 | 4.4 | 3.6 | 3.1 | 2.9 | 2.5 |
| F-test p-value |  | 0.0056 | 0.0020 | 0.0034 | 0.0059 | 0.0109 | 0.0175 | 0.0205 | 0.0348 |
| Log-Likelihood | -112 | -107.8 | -105.2 | -104.4 | -103.8 | -103.5 | -103.0 | -102.2 | -102.0 |
| Partial F-Statistic |  | 8.7 | 5.2 | 1.5 | 1.0 | 0.6 | 0.7 | 1.4 | 0.3 |
| Partial F-test p-value |  | 0.0056 | 0.0291 | 0.2322 | 0.3226 | 0.4480 | 0.4003 | 0.2532 | 0.5961 |
| Partial F-test p-value < 0.05 |  | TRUE | TRUE | FALSE | FALSE | FALSE | FALSE | FALSE | FALSE |

**Table S7**: Linear Regression – Quality of Life (EQ-5D-3L) Not Forced

| EQ-5D-3L | | | | | | | | | |
| --- | --- | --- | --- | --- | --- | --- | --- | --- | --- |
| Rank Included | 0 – forced | 1 | 2 | 3 | 4 | 5 | 6 | 7 | 8 |
| Predictor Added | Constant | Hospital Length of Stay | BMI | Sex | Benzodiazepines and Propofol | Opioids | Age | Days Delirium | Ketamine |
| R^2^ | 0 | 0.31 | 0.38 | 0.42 | 0.43 | 0.44 | 0.44 | 0.45 | 0.45 |
| F-Statistic |  | 16.4 | 11.1 | 8.6 | 6.3 | 5.2 | 4.3 | 3.6 | 3 |
| F-test p-value |  | 0.0003 | 0.0002 | 0.0002 | 0.0006 | 0.0012 | 0.0029 | 0.0064 | 0.0131 |
| Log-Likelihood | -90.2 | -83 | -80.8 | -79.5 | -79.3 | -78.8 | -78.8 | -78.7 | -78.7 |
| Partial F-Statistic |  | 16.4 | 4.3 | 2.6 | 0.2 | 0.9 | 0.1 | 0.1 | 0 |
| Partial F-test p-value |  | 0.0003 | 0.0448 | 0.1188 | 0.6541 | 0.3577 | 0.7405 | 0.7434 | 0.8816 |
| Partial F-test p-value < 0.05 |  | TRUE | TRUE | FALSE | FALSE | FALSE | FALSE | FALSE | FALSE |

**References**

1.	Patanwala AE, Duby J, Waters D, et al.: Opioid Conversions in Acute Care. *Ann Pharmacother* 2007; 41:255–267

2.	Pharmacoeconomic Modeling of Lorazepam, Midazolam, and Propofol for Continuous Sedation in Critically Ill Patients - MacLaren - 2005 - Pharmacotherapy: The Journal of Human Pharmacology and Drug Therapy - Wiley Online Library [Internet]. [cited 2021 Mar 8] Available from: https://accpjournals.onlinelibrary.wiley.com/doi/abs/10.1592/phco.2005.25.10.1319

3.	Guina J, Merrill B: Benzodiazepines II: Waking Up on Sedatives: Providing Optimal Care When Inheriting Benzodiazepine Prescriptions in Transfer Patients [Internet]. *J Clin Med* 2018; 7[cited 2021 Mar 8] Available from: https://www.ncbi.nlm.nih.gov/pmc/articles/PMC5852436/

4.	statsmodels.regression.linear_model.OLS — statsmodels [Internet]. [cited 2021 Mar 5] Available from: https://www.statsmodels.org/stable/generated/statsmodels.regression.linear_model.OLS.html

5.	statsmodels.regression.linear_model.RegressionResults.compare_f_test — statsmodels [Internet]. [cited 2021 Mar 5] Available from: https://www.statsmodels.org/stable/generated/statsmodels.regression.linear_model.RegressionResults.compare_f_test.html
